# Implementation of novel and conventional outbreak control measures in managing a COVID-19 outbreak in a large UK prison

**DOI:** 10.1101/2020.12.07.20234641

**Authors:** Paul C Coleman, Roger Gajraj, Joht Singh Chandan, Anjana Roy, Victoria Lumby, Anna Taylor, Paul Newton, Jonathan Swindells, Éamonn O’Moore

## Abstract

**Background:** SARS-CoV-2 can spread rapidly within correctional facilities. On, following identification of a confirmed COVID-19 case in a prisoner in Prison A (UK), an Outbreak Control Team was convened consisting of prison staff and public health experts from Public Health England and the UK National Health Service.

**Methods:** At the start of the outbreak, four prisoners and 40 staff were isolating with COVID-19 symptoms. An outbreak was declared and full prison lockdown implemented.

Prompt implementation of novel outbreak control measures prevented an explosive prison outbreak, specifically establishment of dedicated isolation and cohorting units, including (i) Reverse Cohorting Units (RCUs) for accommodating new detainees; (ii) Protective Isolation Units (PIUs) for isolating symptomatic prisoners (new detainees and existing residents), and (iii) Shielding Units (SUs) to protect medically vulnerable prisoners.

**Findings:** In total, 120 probable and 25 confirmed cases among prisoners and staff were recorded between. Among prisoners, there were six possible, 79 probable, and three confirmed cases. Among staff, there were 83 possible, 79 probable, and 22 confirmed cases. Testing of symptomatic prisoners was limited for most of the outbreak, with only 33% of probable cases tested. This explains the low number of confirmed cases (three) among prisoners despite the large number of probable cases (n=81; 92%). Over 50% of the initial cases among prisoners were on the two wings associated with the index case.

**Interpretation:** Rapid transmission of SARS-COV-2 was prevented through proactive steps in identifying and isolating infected prisoners (and staff), cohorting new admissions and shielding vulnerable individuals. These novel and cost-effective approaches can be implemented in a wide range of correctional facilities globally and proved effective even in the absence of mass testing.

**Funding Source:** none

**Research in context:** *Evidence before this study:* A systematic literature search on Pubmed from database inception to October 2020 was conducted. The broad terms included were “COVID-19” OR “SARS-COV-2” OR “coronavirus” AND “prison” OR “correctional facility” OR “place of detention” OR “penitentiary” OR “detention centre”. There were no language restrictions. We reviewed reference lists and forward citations of all articles pertinent to the study objectives. We identified 122 results. Only two prison outbreak reports were identified, both focusing on prisons in the USA. Outbreak guidance identified during the literature review was typically adapted from other institutional settings such as care homes and hospital, with a focus on self-isolation, social distancing, reduced admissions and rapid testing. Many of these features form the basis of CDC, WHO and ECDC prison outbreak guidance. However, these measures have not proven effective in many countries, resulting in major and sustained COVID-19 outbreaks in regions such as the USA and South America. Therefore, alternative and cost-effective approaches need to be considered to reduce the transmission of SARS-COV-2.

*Added value of this study:* This study outlines novel outbreak control measures (not reported elsewhere in the literature), that were clearly efficacious in preventing the rapid transmission of SARS-COV-2 in a large UK prison. These include: (1) the timely establishment of “Reverse Cohorting Units” to accommodate new prison admissions, allowing emergent infectious cases to be detected before entering the general population; (2) “Protective Isolation Units” for accommodating confirmed / suspected cases and (3) “Shielding Units” to protect prisoners most at risk from COVID-19.

*Implications of all the available evidence:* The innovative and cost-effective interventions presented here proved effective in controlling the spread of SARS-COV-2, even in the absence of mass testing. The best defence against incursions of future infection into global prisons is implementation of strict outbreak control measures, specifically the screening and quarantining of new prison admissions.

## INTRODUCTION

Severe acute respiratory syndrome coronavirus 2 (SARS-CoV-2) has infected more than 35 million people globally, with over 1 million deaths.^1^ The focus of interest has been the impact of COVID-19 on healthcare and community settings,^2,3^ however, the propensity for explosive COVID-19 outbreaks is of particular concern within prisons and places of detention,^4^ particularly as a second wave of COVID-19 affects many countries.

There have been 42,107 confirmed cases and 510 deaths amongst prisoners in the USA and 540 confirmed cases and 44 deaths among prisoners/probation service users in the UK.^5,6^ Prison populations are at substantial risk of COVID-19 due to staffing,^7^ environmental and host characteristics.^4,8^ Outbreak guidance for prisons from the Centers for Disease Control and Prevention (CDC)^9^ and World Health Organization (WHO)^13^ emphasises the importance of hand-hygiene, social-distancing/isolation, reduced admissions and rapid testing. However, these measures can be difficult to implement^10,11^ particularly against long-term under-investment in European and US prisons.^12^ Other novel and cost-effective approaches are therefore needed.

UK guidance, advised by Public Health England (PHE)^14,15^ outlines specific actions to manage COVID- 19 outbreaks, including the establishment of: (i) Reverse Cohorting Units (RCUs) to accommodate new prison admissions; (ii) Protective Isolation Units (PIUs) for isolating confirmed and suspected cases; and (iii) Shielding Units (SUs) to protect the clinically vulnerable. Here we describe the first prison outbreak in Europe (Prison A) and highlight the vital role of cohorting and isolating in preventing an ‘explosive’ outbreak.

## DESCRIPTION OF THE OUTBREAK

Prison A was notified of a confirmed COVID-19 case in a prisoner (index case) on. The case was tested for SARS-CoV-2 following admission to hospital on. The case did not display COVID-19 symptoms and was discharged on the first Outbreak Control Team meeting was convened (attended by Prison A, PHE public health experts and NHS representatives), four prisoners and 40 staff were isolating with COVID-19 symptoms. An outbreak was declared and prison lockdown implemented (i.e. cessation of all admissions/transfers from courts and other prisons). The outbreak was declared over following on when there had been no confirmed cases among staff or prisoners for over 28 days.

### Establishment of Cohorting and Isolation Units

A SU was established on J-Wing on to safeguard 35 prisoners with chronic medical conditions. Enhanced levels of biosecurity (i.e. measures to prevent introduction of COVID-19) included controlled access via one entrance; full use of PPE by staff; appointment of dedicated prison/healthcare staff and unlocking a maximum of 11 prisoners at one time.

Despite evidence of an on-going outbreak, Prison A reopened to new admissions on due to prison population pressures across England. At this time, an RCU was established with the dual purpose of protecting the main prison population from imported infections, as well as protecting new admissions from any outbreak among the existing prison population. A PIU was also established to isolate confirmed/suspected COVID-19 cases and close contacts for the duration of the infectious period (i.e. seven and 14 days, respectively) (Fig. 1).

**Figure 1.**
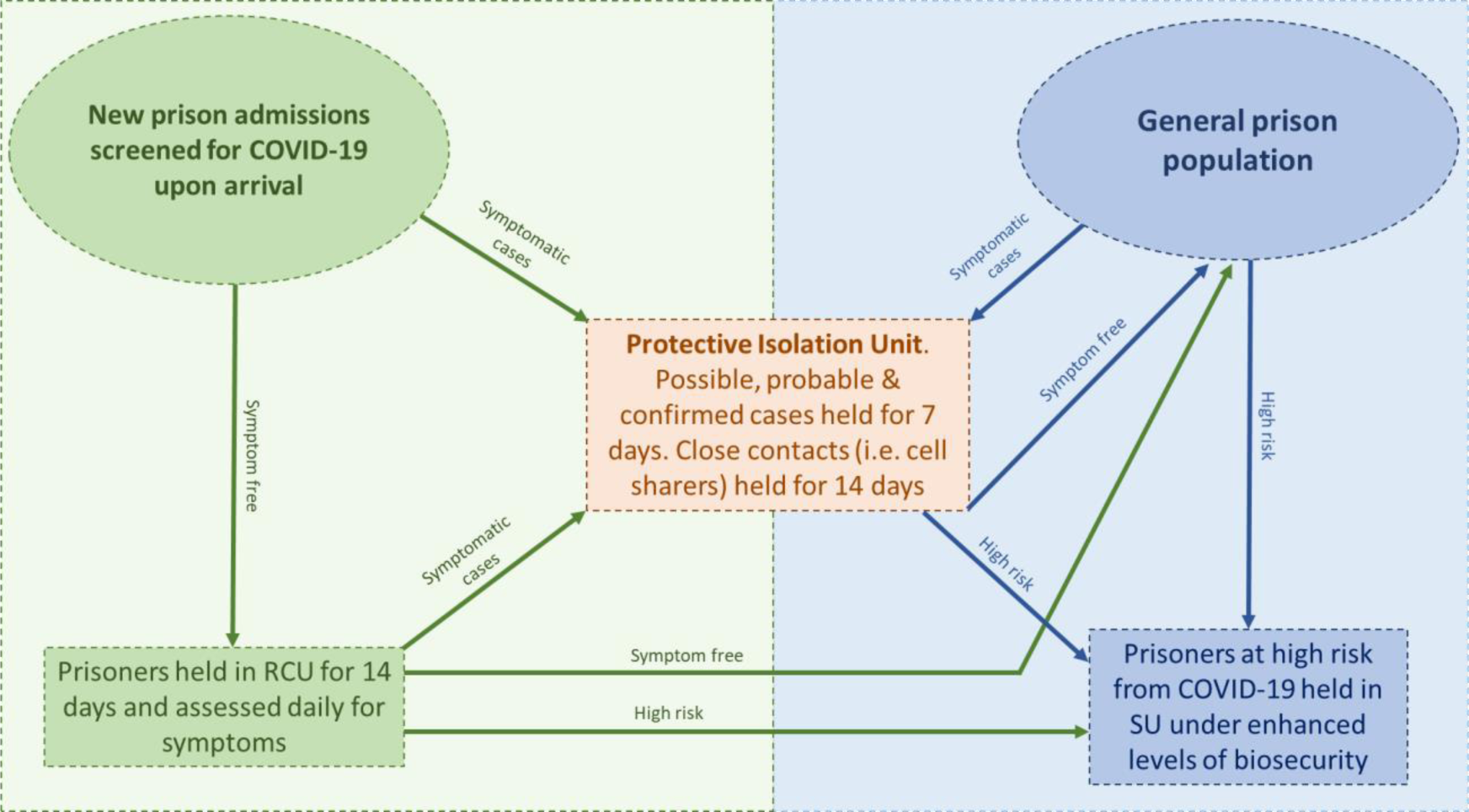
Outbreak control pathway for new admissions and the general prison population in Prison A.

Key features of the outbreak response are the Reverse Cohorting Unit (RCU), Protective Isolation Unit (PIU) and Shielding Unit (SU) From onward, new arrivals underwent healthcare screening for COVID-19 symptoms and were either (i) quarantined in the RCU for 14 days if symptom free, or (ii) held in the PIU for seven days if symptomatic (Fig. 1). Admissions identified as clinically vulnerable to COVID-19 were transferred to the SU once released from the RCU.

Shielding and cohorting units were established in non-central locations, away from the main population, with no thoroughfare and a single point of entry. Single cells were preferred, and facilities (showers, phones, exercise yard etc.) separated from those used by the main population. The Units had designated medical treatments rooms and access to a constant interaction cell (location used for prisoners deemed high risk of suicide). The RCU also had a dedicated area to conduct interviews and clinical assessments.

## METHODS

### Study population

Prison A is a Category B (housing prisoners taken directly from courts) men’s prison in the UK, with a high turnover population and capacity for 977 prisoners. It is composed of seven wings, a social care unit, separation unit and healthcare unit. Each wing accommodates 100–175 prisoners in a combination of single and double occupancy cells. It was holding 950 prisoners and employing 910 staff at the start of the outbreak.

### Case definitions

Definitions of possible and probable cases were made in the context of a confirmed outbreak and therefore deviate from UK national definitions.^14,15)^ Possible cases were individuals reporting symptoms consistent with an upper respiratory tract infection but without COVID-19 symptoms. Probable cases were individuals reporting one or more COVID-19 symptom, i.e. fever or temperature ≥ 37.8°C, new continuous cough or anosmia. Confirmed cases were individuals with a positive SARS-CoV-2 test result.

### Laboratory testing

In the early part of the outbreak, only a minority of symptomatic prisoners were tested because of logistical difficulties in arranging testing undertaken by external healthcare staff. Once infection had been identified in the first cases, those in the prison with symptoms were assumed probable. By, arrangements were established for on-site swabbing, allowing all symptomatic prisoners to be tested. In total, 309 prisoners had been tested by the end of the outbreak), with 294 negative, 13 rejected and two positive results. Five prisoners refused to be tested. Staff sourced testing in the community via local NHS providers.

Prisoners had a throat swab collected by a member of the prison healthcare team using the woven swab from a cobas® PCR Dual or Uni Swab collection kit. These were referred to a local NHS provider and tested using the cobas® SARS-CoV-2 dual target real time PCR assay (Roche Diagnostics, Switzerland). Results were returned electronically to the prison healthcare team.

### Data analysis

An epidemic curve was constructed for staff and prisoners using date of onset of symptoms (symptomatic cases) or date of positive test result (asymptomatic cases). The date of implementation of outbreak control measures are demarcated in the epidemic curve. Demographic and clinical data and SARS-CoV-2 test results were abstracted from prison records by healthcare staff.

The attack rate (AR) was calculated for each prison wing using occupancy data from the start of the outbreak. Prisoner location is defined as the wing the prisoner was located on when symptoms first developed or immediately prior to being tested if asymptomatic.

## RESULTS

Between, 88 prisoners (n = 950, 35.2%) and 184 staff were identified as possible, probable or confirmed COVID-19 cases (Table 1).

**Table 1.** Demographic Characteristics and COVID-19 Diagnosis in Prisoners and Staff at Prison A

|  | **Prisoners**<br>**(N = 88; 9% of**<br>**total prison**<br>**population)** | **Staff**<br>**(N = 184; 20%**<br>**of total staff**<br>**population)** |
| --- | --- | --- |
| **Characteristics** |  |  |
| Age - no. (%) |  |  |
| 20 - 29 | 26 (30) | 32 (17) |
| 30 - 39 | 25 (28) | 23 (13) |
| 40 - 49 | 19 (22) | 37 (20) |
| 50 - 59 | 5 (6) | 34 (18) |
| 60 - 69 | 3 (3) | 3 (2) |
| 70 - 79 | 1 (1) | 0 (0) |
| 80 - 89 | 0 (0) | 0 (0) |
| 90 - 99 | 1 (1) | 0 (0) |
| Unknown | 8 (9) | 55 (30) |
| Sex - no. (%) |  |  |
| Male | 88 (100) | 0 (0) |
| Female | 0 (0) | 0 (0) |
| Unknown | 0 (0) | 184 (100) |
| Hospitalised - no. (%) |  |  |
| Yes | 4 (5) | 1 (0.5) |
| No | 84 (95) | 183 (99.5) |
| **COVID-19 diagnosis - no. (%)** |  |  |
| *Confirmed COVID-19: Total* | 3 (3) | 22 (12) |
| COVID-19 confirmed: Asymptomatic | 1 (1) | 2 (1) |
| COVID-19 confirmed: Atypical symptoms | 0 (0) | 4 (2) |
| COVID-19 confirmed: Typical symptoms | 2 (2) | 16 (9) |
| *Probable COVID-19: Total* | 79 (90) | 79 (43) |
| Probable COVID-19: Not tested | 54 (61) | 61 (33) |
| Probable COVID-19: Tested negative | 25 (28) | 18 (10) |
| *Possible COVID-19: Total* | 6 (7) | 83 (45) |
| Possible COVID-19: Not tested | 4 (5) | 83 (45) |
| Possible COVID-19: Tested negative | 2 (2) | 0 (0) |

### Prisoners

Among prisoners, date of onset of symptoms ranged from (Fig. 2). All three confirmed cases and 22 (35%) of the probable cases occurred in the 29-day period before the PIU and RCU were established on. Over the remainder of the outbreak there were no further confirmed cases and 23 probable cases among the resident population – and 15 probable cases imported from the courts. Four prisoners required hospitalisation. All positive cases were isolated in the PIU as per national guidance.^15^

**Figure 2:**
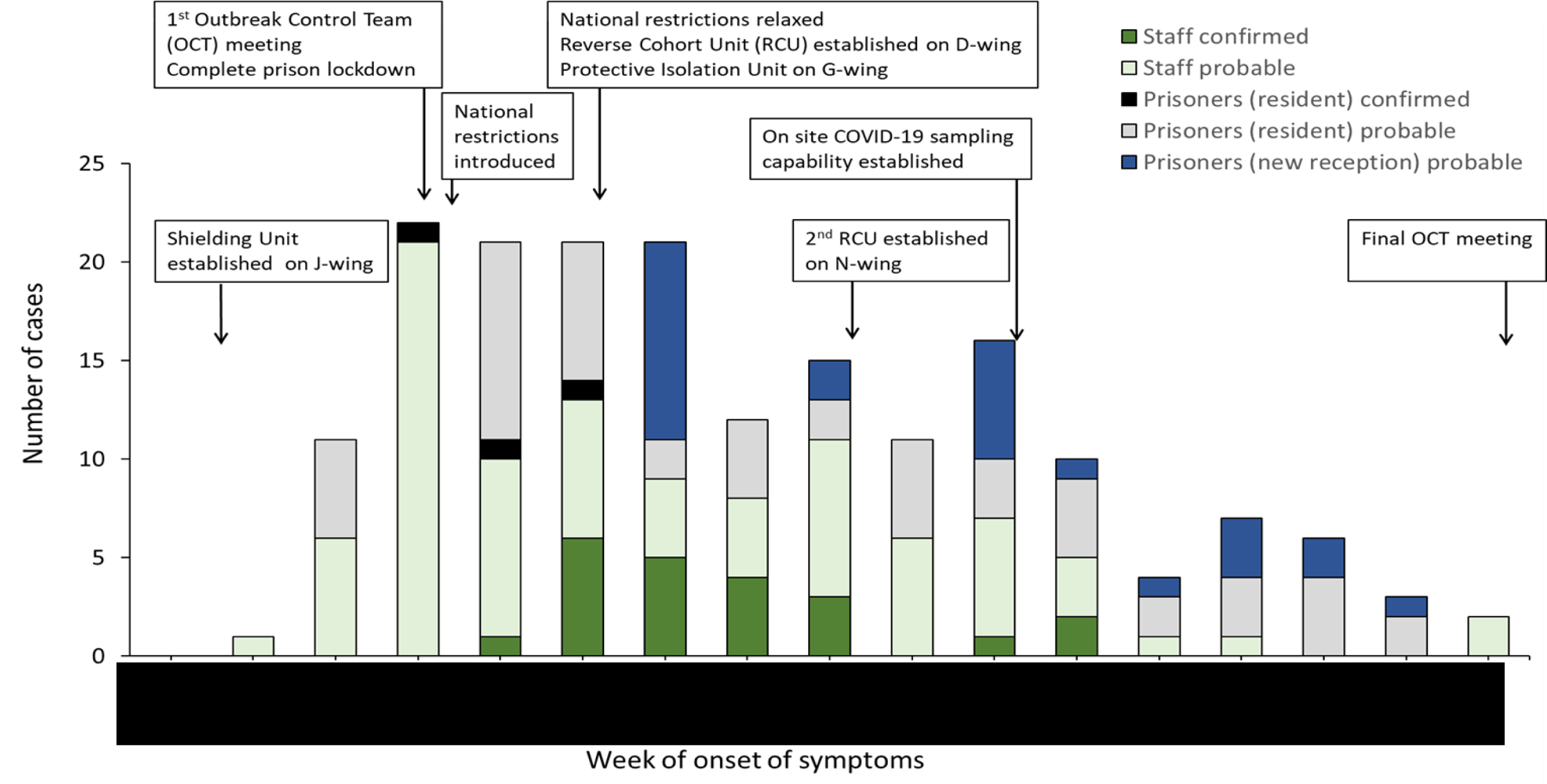
Probable and confirmed cases of COVID-19 by date of onset of symptoms among prisoners and staff linked to Prison A and timeline of key events

All seven wings reported probable or confirmed cases of COVID-19, with at least one case identified on each wing before the PIU and RCU were established on. The attack rate (AR) among prisoners was greatest on D-wing, at 20.0%, followed P-wing (7.8%) and L-wing (5.7%). The attack rate was lowest on K-wing, at 2.9%. Following the PIU and RCU being implemented on the, the majority of new cases were among new admissions to the prison (15; 23.8%).

### Staff

Among staff, there were 83 possible cases, 79 probable cases, and 22 confirmed cases. Two of the 22 confirmed cases had no symptoms and four had atypical symptoms. Only 18 (23%) of the 79 probable staff cases were tested for SARS-CoV-2. Onset of symptoms ranged from. One staff member required hospitalisation (Fig. 2).

## DISCUSSION

This is the first COVID-19 outbreak report in a European prison setting. In the absence of adequate SARS-CoV-2 testing, Prison A was able to control the outbreak through the prompt implementation of novel (RCUs, PIUs and SUs) and standard (social distancing, enhanced cleaning) outbreak control measures. In total, 120 probable and 25 confirmed cases among prisoners and staff were recorded. Here we focus on two main issues that merit further discussion: (i) use of isolation and cohorting units for symptomatic, vulnerable and newly received admissions and (ii) limitations in SARS-CoV-2 testing capabilities.

The WHO have highlighted the importance of increased testing as an essential part of the response to COVID-19. Testing allows effective isolation of true cases and contacts of cases to be identified and quarantined. However, the effectiveness of isolating symptomatic individuals in a COVID-19 outbreak depends on the extent of transmission from pre-symptomatic and asymptomatic infection, and the proportion of cases that are asymptomatic. Data suggests that 20-50% of infections may be asymptomatic^16,17^ and that 40% of transmission occurs before symptoms develop.^18,19^ The infectiousness of asymptomatic individuals is similar to or less than those with symptoms.^20^ Wider testing of individuals with no symptoms therefore allows the identification and isolation of asymptomatic cases who can transmit infection, and identification and quarantining of their contacts. While the original source of infection was likely an asymptomatic staff member moving between wings, as the outbreak progressed it can be assumed that staff continued to be infected from both prisoners and the wider community.

For technical reasons, testing of prisoners was unavailable for most of the outbreak, so only 33% of probable cases were tested. This in part explains the low incidence of confirmed cases (three) among prisoners despite the large number (81; 92%) with classic symptoms. Additionally, there was a delay in Prison A receiving the positive SARS-CoV-2 test result for the index case – subsequently more than 50% of initial cases () were on the wings (G and K) associated with the index case.

The adverse impact on outbreak control of failure to test symptomatic prisoners was mitigated by aggressive identification and isolation of symptomatic prisoners and their contacts. This allowed Prison A to avoid the kind of ‘explosive’ outbreak predicted within prison settings.^4,8^ Even possible cases, without classic COVID-19 symptoms, and their contacts were isolated. It must be noted all members of the prison population (prisoners, staff and visitors) can facilitate the spread of infection. However, a lack of co-ordination between staff and prisoner testing means that the outbreak control team had limited access to staff testing results undertaken by local NHS providers.

Establishment of the RCU and PIU was critical in controlling this outbreak. The RCU allowed Prison A to accommodate new admissions to the prison for a period of 14 days to detect any emergent infectious cases before entering the general population.^21^ Previous literature have confirmed that new admissions are sources of infectious diseases including HIV, Hepatitis B and TB infection.^22^ Data presented here demonstrates that new admissions are an important source of COVID-19. The routine quarantining of new admissions is not advocated by the WHO^13^ or practiced in US prisons, with CDC advising prisons ‘consider’ quarantining ‘if possible’.^9^ This could in part explain the relatively low AR reported for Prison A (6.4%) and other UK prisons^6^ in comparison to the AR of 80% reported for the Marion Correctional Institution^23^ in Ohio and 13% in the Cook County Jail in Illinois, USA.^24^

This investigation has several limitations. Inadequate community and prison testing during the early stages of the outbreak means there is likely an under estimate of confirmed cases among prisoners and staff. As Prison A introduced multiple interventions over a similar timeframe it is difficult to ascertain effectiveness of individual interventions. Due to the nature of the data collected it is not possible to make inferences into behavioural/socio-economic risk factors associated with infection.

In conclusion, In the absence of a vaccine or effective treatment, risks of large outbreaks in prison settings will continue for the foreseeable future. These risks may be escalated as wider community restrictions are relaxed, a second wave of COVID-19 affects many countries and new prison admissions increase as normal criminal justice proceedings resume. The best defence against incursions of future infection into global prisons is implementation of strict outbreak control measures, continued surveillance and testing (specifically the protecting the vulnerable) and, importantly, isolating and cohorting of new prison admissions via approaches such as reverse cohorting unites, protective isolation units and shielding units.

## Data Availability

All data is available upon request from the corresponding author

