## Supplementary material for "Implementation of novel and conventional outbreak control measures in managing a COVID-19 outbreak in a large UK prison": NA

Public Health England

*PHE Research Support and Governance Office*
*Porton Down*
*Salisbury*
*Wilts*
*SP4 0JG*

www.phe.gov.uk

2nd December 2020

For the attention of the medRxiv team

Dear Sir/Madam

**Re: Implementation of novel and conventional outbreak control measures in managing a COVID-19 outbreak in a large UK prison**

The above outbreak report was produced as part of PHE's responsibility to manage the COVID-19 outbreak. As it is not research it falls outside the remit for ethical review.

The report was subject to an internal review by the Research Ethics and Governance Group, which is the PHE ethics committee, and was found to be fully compliant with all regulatory requirements. As no regulatory issues were identified, and ethical review is not required for this type of work, it was decided that a full ethical review would not be needed, and the report was approved by the group.

I hope this has answered your question. Please let me know if you require any further information.

Yours faithfully

E Coates

Dr Elizabeth Coates
Head of Research Governance
Public Health England
